# Irremediable psychiatric suffering in the context of medical assistance in dying: a qualitative study

**DOI:** 10.1101/2021.05.15.21257262

**Authors:** Sisco M.P. van Veen, Andrea M. Ruissen, Aartjan T.F. Beekman, Natalie Evans, Guy A.M. Widdershoven

**Affiliations:** department of Ethics, Law and Humanities at Amsterdam UMC / VUmc & psychiatrist at GGZ inGeest; Haaglanden MC, the Hague, the Netherlands; Amsterdam UMC / VUmc & psychiatrist at GGZ inGeest; Ethics, Law and Humanities at the Amsterdam UMC; department of Ethics, Law and Humanities at Amsterdam UMC / VUmc

## Abstract

**Background:** Establishing irremediability is a central challenge in determining the appropriateness of medical assistance in dying (MAID) for patients with a psychiatric disorder. The objective of this study is to learn how experienced psychiatrists define irremediable psychiatric suffering (IPS) in the context of MAID and what challenges they face while establishing IPS.

**Methods:** In a qualitative study, we performed 11 in-depth interviews with psychiatrists who have experience assessing IPS in the context of MAID.

**Results:** Although determining IPS is, essentially, a prospective assessment, psychiatrists mostly rely on retrospective dimensions, such as the history of failed treatments, when defining IPS. When establishing IPS, psychiatrists face diagnosis and treatment challenges. The main challenge regarding diagnosis is that patients requesting MAID are often diagnosed with more than one psychiatric disorder. Important treatment related challenges are: assessing the quality of past treatments, establishing the limits of approaches that aim to alter the patient’s perception, and managing ‘treatment fatigue’ and treatment refusal.

**Interpretation:** IPS is mostly defined in terms of failed past treatments. Challenges regarding diagnosis and treatment complicate the process of establishing IPS. The finding that most patients are diagnosed with more than one psychiatric disorder calls for a critical appraisal of the due diligence procedure for MAID. In response to the challenges related to treatment, intersubjective clinical criteria for IPS in the context of MAID should be developed. Also, further investigation of the concept of ‘treatment fatigue’ in psychiatry may provide insight into why patients requesting MAID.

**Registration:** this study was preregistered under osf.io/2jrnd.

## Introduction

Medical assistance in dying (MAID), also known as physician assisted death, has been legalized in an increasing number of jurisdictions around the world. (1) In 2023, Canada will join a small group of countries that allow MAID for *persons with a psychiatric disorder* (PPD). (2) In the Netherlands, MAID is allowed for irremediable psychiatric suffering (IPS) since the 1990s, and the last decade has seen a marked increase in cases. In 2020 MAID was performed 88 times for psychiatric suffering (1.3% of all MAID cases). (3) The number of MAID requests by patients with a psychiatric disorder is much higher, but over 90% of these requests are denied. (4)

The main legal requirements in the Netherlands are that the patient must be able to make a competent request, that the patient’s suffering must be unbearable and irremediable, and that the patient and physician have to agree that there are no other reasonable treatment options. Also, an assessment by an independent physician and, in case of psychiatric suffering, a third assessment by an independent psychiatrist, preferably with specific expertise regarding the patient’s disorder, are required. (4)

While there are concerns about decision-making ability, the central dilemma of MAID for PPD appears to revolve around applying the concept of irremediability to psychiatric disorders. (5) A recent scoping review identified a multitude of conceptual articles addressing irremediability in the context of psychiatric MAID, but few empirical studies. (1)

Surveys estimate that 46% of Dutch psychiatrists received an explicit MAID request at least once in their career, and 4% actually assisted in the death of a PPD. (4) The experiences of psychiatrists who have handled MAID requests can be seen as an important source of knowledge about the challenges of establishing IPS in practice. The aim of this study is to learn how experienced psychiatrists define psychiatric suffering as irremediable in the context of a MAID request, and what challenges they face while establishing IPS.

## Methods

### Participant selection

The participating psychiatrists were purposefully sampled from the authors’ professional networks. Psychiatrists who performed an independent consultation during a MAID-procedure and psychiatrists who performed MAID themselves were included. Both known proponents, known opponents and psychiatrists with moderate views on MAID for PPD were actively invited to ensure the inclusion of a range of perspectives.

### Data collection

A topic list was assembled based on the clinical and ethical experience of the authors. Additional items were identified through a simultaneously ongoing systematic review of relevant literature. (1) The topic list was piloted on two senior psychiatry residents. After interview five and seven, the topic list was slightly revised, based on interim-analysis and discussion amongst the authors.

Participants were interviewed at their place of work or at a place of their choice. During the interviews, only the participant and interviewer (SvV) were present. Field notes were made during the interviews. All interviews were audio-recorded and subsequently transcribed.

### Data analysis

The method of analysis was the Grounded Theory approach, as developed by Charmaz (2006). (7) All data were entered into MAXQDA-software, version 2018.2. Coding was performed by SvV and AR. The codes were compared and disagreements were resolved through discussion, and attention was given to similar and divergent cases. Coding followed three consecutive stages. In the open coding stage, a set of codes was created describing elements psychiatrists incorporated in their decision about IPS and challenges they faced when doing so. During the second, ‘focused coding’ stage, the codes were organized into potential themes, and overlap between categories was minimized. The themes were reviewed by the authors and used for revision of the topic list. In the third, axial coding phase, the relationships among and patterns between the various themes were examined, after which the over-arching themes were refined and the final categories formulated. After 11 interviews theoretical saturation was reached. During the last four interviews, with participants from various backgrounds, no new themes emerged.

### Ethical approval and informed consent

This study was approved by the medical ethics committee of the VU Medical Centre Amsterdam (2018.661). Participants received an information letter via mail and written informed consent was obtained.

### Quality criteria

For reporting, the Consolidated criteria for reporting qualitative research (COREQ) were followed. (8) The study-protocol was preregistered, during data-analysis, at the Open Science Framework under osf.io/2jrnd.

## Results

Eleven psychiatrists were included in the study, their characteristics are described in Table 1. Initially 17 psychiatrists were invited; five did not have the required experience with MAID and one did not respond. The clinical areas of expertise of the 11 psychiatrists included: anxiety disorders, autism spectrum disorders, bipolar disorders, depressive disorders, minor mental disability, obsessive-compulsive disorders, personality disorders, severe psychiatric illness, and somatic symptom disorders. The sample contained both proponents, opponents and psychiatrists with moderate views on MAID. The mean interview duration was 62 minutes (range 48-77 minutes). The analysis focused on two main questions: the definition of IPS and the challenges psychiatrists face when establishing irremediability.

**Table 1:** participant characteristics.

| **Table 1: participant characteristics** |  |
| --- | --- |
| Mean age (range) | 55 (35-64) |
| Female / Male | 7 / 4 |
| **Place of work** |  |
| Specialized mental health clinic | 5 |
| Academic hospital | 2 |
| General hospital | 3 |
| Expertise Centre Euthanasia | 1 |
| **Experience with MAID** |  |
| Independent expert | 10 |
| Performed MAID* | 2 |
| **Religious background** |  |
| Non-believer | 4 |
| Christian upbringing, non-practicing | 5 |
| Practicing Christian | 1 |
| Missing | 1 |
| MAID = Medical Assistance in Dying |  |
| *One participant both performed MAID and functioned as an independent expert (in separate procedures). |  |

### Definition of IPS

Irremediability, in general, is a *prospective* concept because the physician has to make a judgement about the future course of a disease. Establishing irremediability is therefore seen as synonymous to establishing a poor prognosis.

> *“When someone says: I have a chronic disorder that will not go away, when all the experts agree and when it causes suffering, then the suffering is irremediable” - P6*

Various participants argue that making meaningful prognostic claims about psychiatric suffering is challenging and some fear that it is impossible. In part, because the psychiatrist has to make a claim about a patient who potentially has decades to life.

> *“We all know examples of people with, for example, therapy-resistant depression, which we have more or less given up (*…*) and then a few years later you find out to your surprise that they have found their way and have recovered. That makes [establishing IPS] very difficult*.*” - P11*

Given the limits of prospectively establishing IPS, the participants emphasize *retrospective* dimensions. In particular, participants state that IPS is dependent on the treatment history and underline that substantial attempts to reduce suffering should have been tried and failed.

> *“If someone has gone through all the [treatments] and there is nothing left of which you can say: if you do that, it will be different. (*…*) Then I think [the suffering] is irremediable*.*” – P5*

Because of the complex nature of psychiatric suffering and treatment, most participants see uncertainty as unavoidable. Comparing psychiatric suffering to somatic suffering, participants mention that establishing irremediability for the latter is more certain due to the clear biological substrate of the disease and also because patients will often die from these diseases unassisted.

> *“I think [in psychiatry] it is very complicated [to establish irremediability], with cancer you just know. Chemotherapy does nothing, there are no other options and then the tumor starts to grow and then… it just stops*.*” - P2*

A consequence of this uncertainty is that it leaves room for substantial interprofessional differences. As one participant, somewhat hyperbolically, puts it:

> *“I think that if the same patient is seen by 10 different psychiatrists, you will get 10 completely different letters [describing the patient and advising on irremediability]*.*” – P7*

Another participant, however, argues that uncertainty should be accepted:

> *“We just have to accept that there will always be some degree of uncertainty. The moment I, as an independent psychiatrist, say, ‘I think the legal criteria have been met’, then there is an uncertainty. There is a confidence interval around it. (…) Because it concerns a dichotomous choice of life or death, we want absolute 100% certainty. (…) But this is not possible*.*”- P8*

### Challenges in establishing IPS

In relation to establishing IPS, participants mention challenges regarding diagnosis and treatment.

#### Challenges regarding diagnosis

Participants say that patients who request MAID are often diagnosed with more than one psychiatric disorder, which complicates the assessment procedure and the application of evidence-based guidelines. One participant argues that, therefore, a second opinion by an all-round psychiatrist is the best way to determine IPS.

> *“Most people who request MAID on psychiatric grounds do not suffer from one disorder. [Take] a patient who complains most about depression, but she is also an adolescent, she is traumatized (…) she has psychosomatic complaints and there are systemic problems due to a symbiotic relationship between mother and daughter. (…) What expert is best equipped to independently assess this patient [and come to a conclusion about IPS]? I think that it is better to have a more generalized perspective in this case, than focus on one specific disorder. – P10*

It may also happen that the independent psychiatrist comes to a different diagnosis in the course of the MAID procedure, often leading to new treatment options. P1 describes a patient he diagnosed with a new disorder, but also nuanced the accuracy and clinical importance of these new diagnostic insights:

> *“Recently [I saw] a woman with a very serious social phobia, who was completely stuck in her life (…) [after additional diagnosis] she turned out to be autistic (…) But then I immediately think: this [new diagnosis] is just a conclusion from a number of questionnaires or interviews. So, I am not exactly sure what the value is of such a new diagnosis. - P1*

#### Challenges regarding treatment

Most challenges in establishing IPS are related to treatment. For instance, it may be challenging to assess the quality of past treatments, especially when it concerns psychotherapy. In order to evaluate past treatments, participants review the patient’s file, consider the reputation of the center where the patient was treated and take into account the patient’s view on therapy. One participant describes this process as follows:

> “*You look at goals that have been set; have they been achieved? Was there enough commitment? Was the patient motivated? You will also try to understand the content of the therapy and ask the patient about this as well*.*” – P9*

When evaluating past attempts to reduce suffering, participants regard the relevant treatment guidelines as important resources.

> *“For example, when assessing someone with a mood disorder, I want to read in the correspondence or hear from the patient that the usual steps in the guidelines have been followed*.*” - P8*

Participants also emphasize the value of interventions that try to diminish suffering by altering the patient’s perception of the complaints. Acceptance and commitment therapy is often mentioned in this context, but participants also refer to recovery-based or rehabilitation-based approaches or ‘using the handicap model’.

> *“Those kinds of therapies give a different dimension to the patient’s experience. [We have to try to help patients to accept] that everything will not go back to the old level of the past, when they were not yet ill and everything was still possible, and start a new phase of life with limitations*.*” - P9*

Based on this view of psychiatric treatment, some participants argue that the options are practically endless. Therefore, irremediability cannot be established.

> *“In psychiatry, it is almost never the case that there are no treatment options at all, you can also give recovery-based care, or supportive care, or long-term clinical care with daytime activities. I mean, there is always some form of care possible. Because people usually do not die from it*.*” – P10*

In contrast, other participants feel that endlessly working towards acceptance or recovery is not a reasonable answer to patient’s request for MAID.

> *It is almost never possible to predict anything in psychiatry. (…) And at the same time, I also think it is a bit cowardly to keep saying that [there are always treatment options], because that gets you nowhere. (…) I think ultimately you don’t help people with this point of view. - P7*

Various participants noticed ‘treatment fatigue’ when discussing new treatment options with the patients. The participants suggest that this fatigue may be due to the typically long treatment history of patients requesting MAID.

> *“He was just tired, he was fed up with it, he thought [starting a new treatment] made no sense at all” - P1*

Sometimes, within the current legal framework, additional treatment is necessary in order to be eligible for MAID. As a result, patients may try a treatment just ‘to check the box’. Various participants doubt whether treating unmotivated patients under these conditions is effective, especially when it concerns psychotherapy. Participants mention that there is no research available about the efficacy of treatment with unmotivated patients.

> *“Our evidence-based guidelines are based on people who wanted to be treated, it has never been shown that a treatment can be effective if someone does not want it at all. Regardless of whether it is practically feasible. So, I think there is a great tension there that our profession has no answer to*.*” - P4*

One participant emphasizes that there are also cases where the outlook of MAID can increase treatment motivation.

> *“MAID can turn out to be a real possibility if it all doesn’t work. And if you, as a patient, dare to trust that this possibility [MAID] is there at the end of the tunnel, then you may continue that tunnel for a bit longer*.*” - P10*

A final challenge regarding treatment is refusal of new treatments by the patient. Many participants try to determine whether the refusal is reasonable. If not, they will not see suffering as irremediable, and stop the MAID-review-procedure.

> *“If there are realistic treatment options that can be tried within a reasonable period of time, and someone refuses, I think it is also reasonable that the [MAID]-procedure should stop”. - P4*

Several participants conceptualize reasonability in terms of a balance between burden and possible benefit. The treatment history is also relevant. One participant says:

> “*I think it is a matter of balancing. What treatment options are there? And what should someone do for that? And does that then outweigh any expected effect? And after how long can you expect that? And you also take into account someone’s treatment history; is someone still susceptible to change?” – P1*

## Interpretation

This study provides a first exploration of how psychiatrists define IPS in the context of MAID and what challenges they encounter while establishing IPS. Although IPS is seen as a prospective concept, participants mostly refer to retrospective dimensions and more specifically to the failure of past treatments, when defining IPS. This appears logical, for various studies have shown that failed past treatments are a predictive factor for chronicity in psychiatry (9–11). This study also confirms earlier findings that psychiatrists struggle with uncertainty as a distinctive element of the definition of IPS. In the conceptual debate about IPS in the context of MAID, much attention is given to uncertainty, and it is often used as an argument against MAID for PPD. (1,12) Yet, most participants feel that refusing MAID due to uncertainty alone does not do justice to the individual patient’s request for MAID. We have to acknowledge that absolute certainty about the prognosis of any type of suffering is epistemologically impossible. We should therefore also aim to find a reasonable balance between the need for certainty and the need to assist individual patients who wish their suffering to be ended. (13)

Participants mention differences in professional opinion when establishing IPS. An earlier casefile study of PPD that died through MAID showed that there was disagreement between physicians about IPS in 11% of cases. (14) Although the number in this earlier study is relatively low, differences in professional opinion do raise questions about the clarity of the concept of IPS and the potential for a high degree of subjectivity in its use in clinical settings. Further understanding and higher levels of unicity could be achieved by establishing intersubjective clinical criteria for IPS in the context of MAID.

When establishing IPS, participants face different challenges regarding diagnosis. The participants mentioned that most patients who request MAID are diagnosed with more than one psychiatric disorder. This finding is in line with earlier studies reporting that 71-79% of psychiatric patients that died through MAID in the Netherlands were classified with more than one psychiatric disorder. (14–16) This raises questions about whether all distinct treatment protocols for each disorder must be followed completely before IPS can be established. Also, it raises questions about the current Dutch guideline which states that an independent psychiatrist ‘with specific expertise about the patient’s disorder’ should be consulted. In practice it may prove difficult to identify which expert is best suited, and, as one participant mentioned, an independent assessment by a psychiatrist with a more generalized view might be more suitable. Another question is whether new diagnoses during the MAID-procedure should lead to new treatment plans. A recent case-report has shown that this could lead to recovery in individual cases. (17)

The participants also describe challenges regarding treatment when establishing IPS in the context of MAID. They mention that it can be difficult to evaluate the quality of earlier treatments. They also struggle to find a reasonable limit to demanding more interventions that try to diminish suffering by altering the patient’s perception of the complaints. Also, participants recognize a form of ‘treatment fatigue’ among PPD requesting MAID, due to their often long treatment history. The term ‘treatment fatigue’ has been researched in the context of HIV and type 1 diabetes, but it has not yet received attention in psychiatry. (18) Research on treatment fatigue in psychiatry could provide new insights and might result in better care for patients with treatment resistant psychiatric disorders. Eventually this may lead to subsidiary alternatives to MAID.

The participants also mentioned struggling with treatment refusal while establishing IPS. The finding that treatment refusal is a relevant issue is in line with earlier studies, showing that 56% of PPD that received MAID have refused some form of treatment. (15) It also justifies the considerable attention treatment refusal has received in the conceptual literature about IPS in the context of MAID. (1) We have seen that psychiatrists largely base their assessment of IPS on failed treatments. Therefore, it is reasonable to assert that when a patient refuses a substantial number of treatments, this hampers the opportunity to properly asses irremediability. In our view, a process of shared decision-making is most suited to decide on the limits of treatment refusal. (13) The patient should be aware of the potential benefits and burdens of new treatments and the psychiatrist should try to understand why the patient refuses certain treatments. Intersubjective clinical criteria for IPS in the context of MAID could help to establish reasonable limits for treatment refusal.

The main limitation of this article is the limited number of psychiatrists interviewed. The main strength of the study is that it adds in-depth empirical knowledge to a controversial topic that is gaining global significance and that is subject of a long-lasting conceptual debate. Finally, future research should also focus on the patient’s views on IPS in the context of MAID. (6)

In conclusion, establishing irremediability of suffering is a central challenge in the context of MAID for PPD. We performed 11 in-depth interviews with psychiatrists who have experience assessing IPS in the context of MAID to learn how they define IPS and what challenges they face while establishing IPS. Although IPS essentially is a prospective concept, psychiatrists mostly refer to retrospective dimensions when defining IPS. More specifically, they mainly focus on the history of failed treatments. When establishing IPS the psychiatrists face challenges related to both diagnosis and treatment. The main challenge regarding diagnosis is that patients requesting MAID are mostly diagnosed with more than one psychiatric disorder. Important treatment related challenges are: assessing the quality of past treatments, establishing the limits of approaches that try to diminish suffering by altering the patient’s perception, and managing treatment refusal. In response to these challenges, a better understanding of ‘treatment fatigue’ among PPD might result in better care for patients with treatment resistant psychiatric disorders, may yield a better understanding of treatment refusal, and may eventually lead to subsidiary alternatives to MAID. Finally, by drafting intersubjective clinical criteria for IPS in the context of MAID, many of the identified challenges can be mitigated.

## Data Availability

The qualitative data (interviews) and coding is available upon request (in Dutch).

